# Childhood Bullying as a Risk Factor for Late-Life Psychological Distress and Cognitive Impairment

**DOI:** 10.1101/2023.09.04.23295046

**Authors:** Iván Mejía-Guevara, V.S. Periyakoil

## Abstract

In the United States, non-Hispanic Black (19%) older adults are more likely to develop dementia than White older adults (10%). As genetics alone cannot account for these differences, the impact of historical social factors is considered. This study examined whether childhood and late-life psychological distress associated with dementia risk could explain part of these disparities. Using longitudinal data from 379 White and 141 Black respondents from the Panel Study of Income Dynamics, we assessed the association between childhood bullying and late-life dementia risk, testing for mediation effects from late-life psychological distress. Mediation analysis was computed via negative binomial regression modeling, stratified by race (White/Black), type of bullying experience (target, bully, and bully-target), and the age range at which the experience occurred (6-12, 13-16). The results indicated that late-life psychological distress fully mediated the association between Black respondents who were bullies and dementia risk. However, no significant association was observed among White respondents. These results suggest that interventions aimed at preventing and treating psychological distress throughout the lifespan could be crucial in mitigating the development and progression of dementia risk.

## INTRODUCTION

In the United States, Black older adults (aged 65+) are twice as likely to develop dementia compared to White older adults (10%). While some research suggests that genetic risk factors, such as Apolipoprotein E-4 (APOE-4), may differ by race, these differences are unable to account for the significant differences in dementia prevalence that have been observed.^1–3^ Moreover, diagnosis of dementia in Black older adults is often missed or delayed, meaning that these disparities are likely more severe than currently suggested by the literature.^4,5^ Because race and ethnicity are social constructs with little genetic basis, it has been hypothesized that historically perpetuated environmental and sociopolitical factors may instead explain differences in dementia disparities.^6–9^ The goal of this study was to examine the association between childhood bullying and late-life psychological distress and dementia.

Stress is associated with the development and progression of neurogenerative diseases and mental disorders.^10,11^ Stress acts in large part through the activation of the Hypothalamic-Pituitary-Adrenal, which elevates corticosteroid levels,^12,13^ and it has been hypothesized that a “Vicious Cycle of Stress” exists in which stress causes disease and disease causes stress, accelerating and producing neuropsychiatric complications—including depression, anxiety, and aggressive behavior.

Childhood bullying,—usually defined as the use of physical or emotional power to control or harm others—a marker of chronic stress, has been associated with severe adult physical and mental health problems.^14–16^ For instance, victimization has been linked to higher levels of low-grade systemic inflammation in adulthood than non-victims.^14^ Depending on the type of bullying, targets (or victims of bullying) or perpetrators may internalize^17^ (e.g., depression, anxiety, social isolation) or externalize^18^ (e.g., physical/relational aggression, defiance) behavior problems. Previous research indicates that compared to other ethnicities, Black adolescents have higher rates of both perpetrating and experiencing bullying, and those who experience any bullying are more prone to internalizing symptoms than those who do not.^19,20^

Childhood stress and stress during adulthood are known to be risk factors for cognitive decline, Mild Cognitive Impairment (MCI), or dementia in late life.^21–23^ A previous study found that old-age Black people showed less risk of emotional distress compared to Non-Hispanic White people.^24^ However, a recent study revealed that serious psychological distress (SPD) is linked to Alzheimer’s disease and related dementias (ADRD), with Black people having greater odds of ADRD than Non-Hispanic White people. The study also found that SPD plays a significant role in driving racial disparities in ADRD prevalence.^25^

This study examines whether childhood bullying increases susceptibility to late-life dementia and differential responses among various racial groups and whether late-life emotional distress may mediate that potential association.

## METHODS

Data for this study are from the Panel Study of Income Dynamics (PSID)—the world’s longest-running national household panel survey that collects longitudinal data on measures of economic and social well-being. Its sample size has increased from approximately 5,000 families in 1968 to over 9,000 families in 2019, and it currently has over 50 years of data collected on the same families and their descendants, creating a robust cornerstone for empirically-based social science research.^26^ In this study, we used several instrument measures to examine the impact of childhood/adolescence adverse event exposure on the risk of developing dementia later in life.^27^

### Outcome, mediator, and exposure

#### Outcome: AD8 Dementia Screen

The AD8 instrument was added by PSID for the first time in 2017 as a brief instrument to detect mild cognitive impairment (MCI) and to help distinguish between normal aging and individuals at risk for developing ADRD.^28^ Combined with the longitudinally of the PSID, this confers an unprecedented opportunity to examine the effects of social factors in early life that may contribute to dementia risk. Although dementia risk is not directly assessed in PSID, AD8 is a reliable alternative with good sensitivity (>84%) and specificity (>80%) and highly correlated (r=0.75) with the Clinical Dementia Rating (CDR), a gold standard global dementia rating system.^29,30^ AD8 is a short instrument composed of eight items that take, on average, only 3 minutes to complete. It has been validated as an informant and self-reported instrument and is well-suited for PSID data because only one person is interviewed per family.^29^ **Table 1** lists the eight response items of the AD8 instrument.

**TABLE 1.** Survey instruments to assess psychological distress (K6) and dementia risk (AD8) that were included in the PSID.

| **K6 non-specific distress scale** |  | **AD8 Dementia screening interview** |  |
| --- | --- | --- | --- |
| During the past 30 days, how often did you feel...?* |  | Items ** |  |
| a. nervous |  | a. Problems with judgment |  |
| b. hopeless |  | b. Less interest in hobbies/activities |  |
| c. Restless or fidgety |  | c. Repeats the same things over and over |  |
| d. so depressed that nothing could cheer you up |  | d. Trouble learning how to use a tool, appliance, or gadget |  |
| e. that everything was an effort |  | e. Forgets correct month or year |  |
| f. worthless |  | f. Trouble handling complicated financial affairs |  |
|  |  | g. Troubling remembering appointments |  |
|  |  | h. Daily problems thinking and/or memory |  |
| Mean (SD) |  |  |  |
| White | Black | White | Black |
| 10.99 (3.99) | 10.94 (6.42) | 1.09 (1.79) | 1.15 (2.62) |

\* Based on a 5-point Likert scale: 1) none, 2) a little, 3) some, 4) most, 5) all of the time.
\*\* Yes (a change), No (no change), N/A (don't know).

#### Mediator: K6 Emotional Distress Scale

Psychological distress was examined as the primary mediator, which was assessed using the Kessler Psychological Distress Scale (K6). The K6 instrument design distinguishes cases of serious mental illness from non-cases and demonstrates good specificity and sensitivity.^31^ The 2016 PSID Wellbeing and Daily Life Supplement of the PSID (PSID-WB) included the six items of the K6 instrument to measure respondent’s positive and negative emotions in the last 30 days using a 5-point Likert scale that we inverse-recoded as 1 (None of the time), 2 (A little of the time), 3 (Some of the time), 4 (Most of the time), and 5 (All of the time).^32^ The K6 was scored using an unweighted sum of the answer responses (**Table 1**).

#### Exposure: Childhood Bullying

We examined childhood bullying experiences as the primary exposure. Retrospective data for respondents 6 to 16 years old included four items addressing peer victimization and perpetration. The data are publicly available in the Childhood Retrospective Circumstances Study 2014 (PSID-CRCS).^33^ The four items consisted of the information reported separately for ages 6-12 and 13-16 and whether the event occurred in or out of school. All items are based on a 4-point Likert scale with four extreme ordinal categorical responses: 1) A lot, 2) Sometimes, 3) Rarely, and 4) Never. We recoded indicators as binary experiences, with values of 1-2 (a lot or sometimes) recoded as 1, indicating moderate or high exposure, while values 3-4 (rarely or never) were recoded as 0, indicating rare or shallow exposure. To better assess child bullying, we defined three types of experiences:^34^ ‘target’, ‘bully’, and ‘bully-target’. A target is a person repeatedly exposed to unwanted, aggressive behavior by her peers; a bully consistently causes emotional, physical, or social harm to peers; and a bully-target describes an individual who was both a bully and was bullied. To create the bully-target category, we combined responses from individuals who had experienced both sides of bullying—as targets and perpetrators.

#### Covariates

We adjusted all the statistical models for sex, the number of years of schooling, and race, which are essential demographic characteristics related to stress or dementia in previous studies.^35^ Sex was binary, and the number of years of schooling was recoded as a categorical variable: 1 for less than 12 years, 2 for 12 years completed, 3 for 13-15 years, and 4 for 16-19. Finally, we assessed race using two separate items reported in the 2017 family PSID file, which initially included six categories (White, Black/African American (henceforth “Black”), American Indian or Alaska Native, Asian, Native Hawaiian or Pacific Islander, and Other). However, we only kept non-Hispanic White and non-Hispanic Black respondents because the small sample from the other racial groups impeded a robust statistical analysis. To exclude other racial groups, we used a single ‘Hispanic’ category from another item reported as Hispanic descent, represented by seven different ethnicities (Mexican, Mexican American, Chicano, Puerto Rican, Cuban, and Other Spanish/Hispanic/Latino) after we combined recoded race and Hispanic variables to obtain the two final categories, ‘non-Hispanic White’ and ‘non-Hispanic Black’—hereinafter referred to as White and Black for simplicity, respectively. We conducted stratified analyses using this resulting variable to examine race/ethnic-related variation.

#### Study Population

We linked the PSID-Wellbeing and Daily Life (WB) and PSID-Childhood Retrospective Circumstances Study (CRCS) supplements to the 2017 wave of the PSID individual and family files to measure the association between early-life exposure to bullying and late-life psychological distress (K6 instrument) and dementia risk (AD8 instrument). The 2017 PSID family roster included 9,607 families/respondents (1,038 Hispanic/Latino, 4,460 Non-Hispanic White, 3,402 Non-Hispanic Black, and 707 Non-Hispanic other or missing). For this study, we focused on the 1,461 respondents aged 65 and older. We next excluded the following respondents: 183 identified with a race/ethnicity group other than Non-Hispanic White/Black, 485 with no linkable information from the 2014 PSID-CRCS, and 270 who were not eligible for screening of dementia risk in 2017 PSID-WB. The final analytic sample included 523 respondents comprised of 141 Non-Hispanic Black and 379 Non-Hispanic White respondents (**Figure 1**).

**Figure 1.**
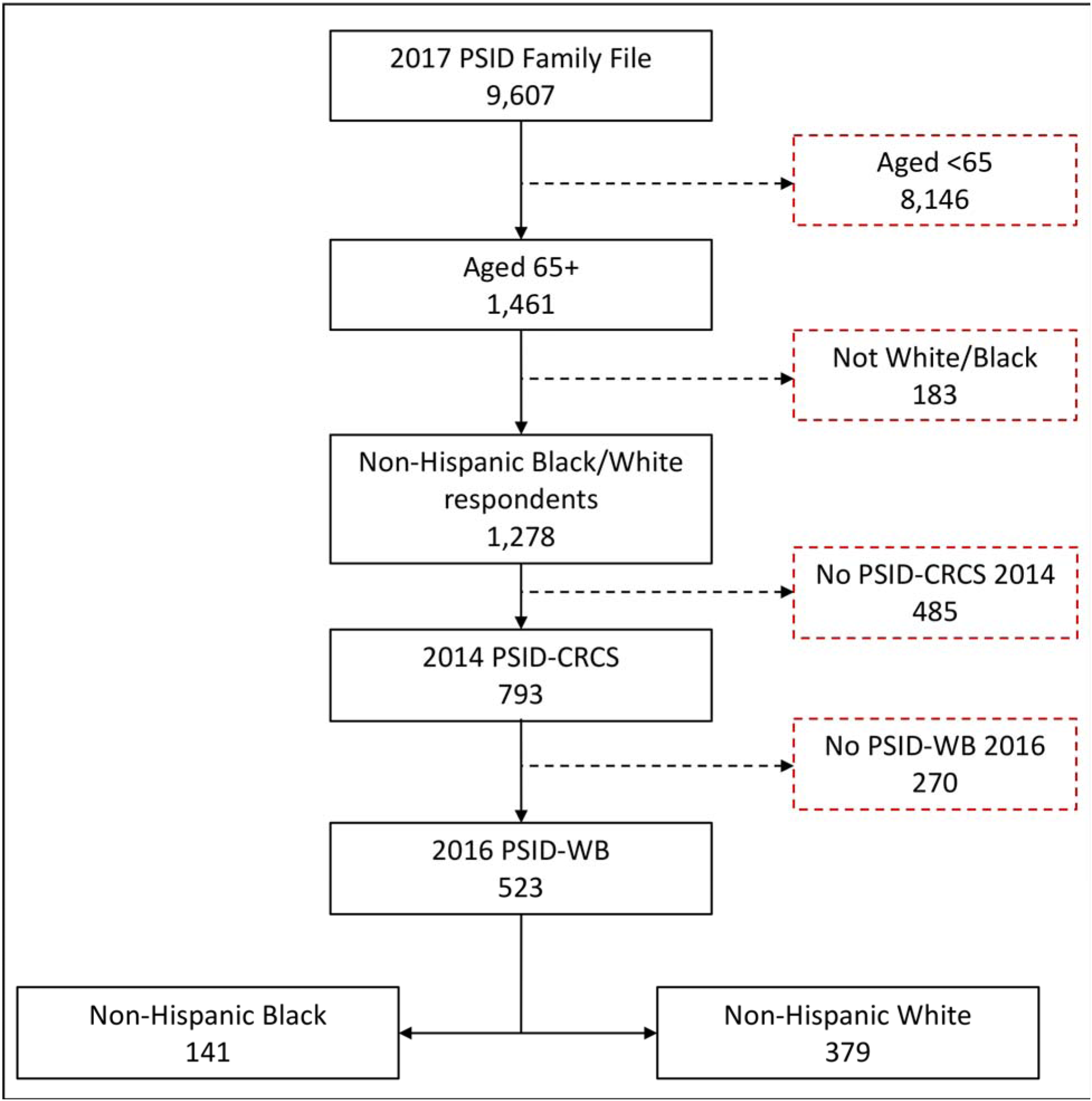
Final analytic sample for the mediation analysis of childhood bullying, psychological distress (H6), and dementia risk (AD8) by race. We merged the PSID family and individual files (2017), the Childhood Retrospective Circumstances Study (CRCS 2014), and the Wellbeing and Daily Life supplement (WB, 2017) files. Respondents were excluded if they were younger than 65 in 2017 (8,146 respondents), not non-Hispanic White or Black (183 respondents), did not participate in the CRCS 2014 (485 respondents), and were not eligible for screening of dementia risk in the 2017 PSID-WB (270 respondents).

### Statistical Mediation Analysis

We followed a traditional approach for mediation analysis to assess the association of childhood bullying and late-life (age 65+) increased risk of dementia,^36,37^ mediated by late-life psychological distress. **Figure 2** illustrates the schema of the mediation analysis and the specifications of the three corresponding models. The first model calculates the total effect of the long-term association between childhood *Bullying* and increased risk of dementia (AD8) while adjusting for individual demographic characteristics. The second model tests the long-term association between childhood *Bullying* and late-life psychological distress (K6). The third model assesses the association between Bullying and AD8, including the potential mediator K6. We used Negative Binomial rather than Poisson regression models due to evidence of overdispersion in the distribution of AD8 (e.g., mean=1.15 and SD=2.62 for Black respondents, **Table 1**).

**Figure 2.**
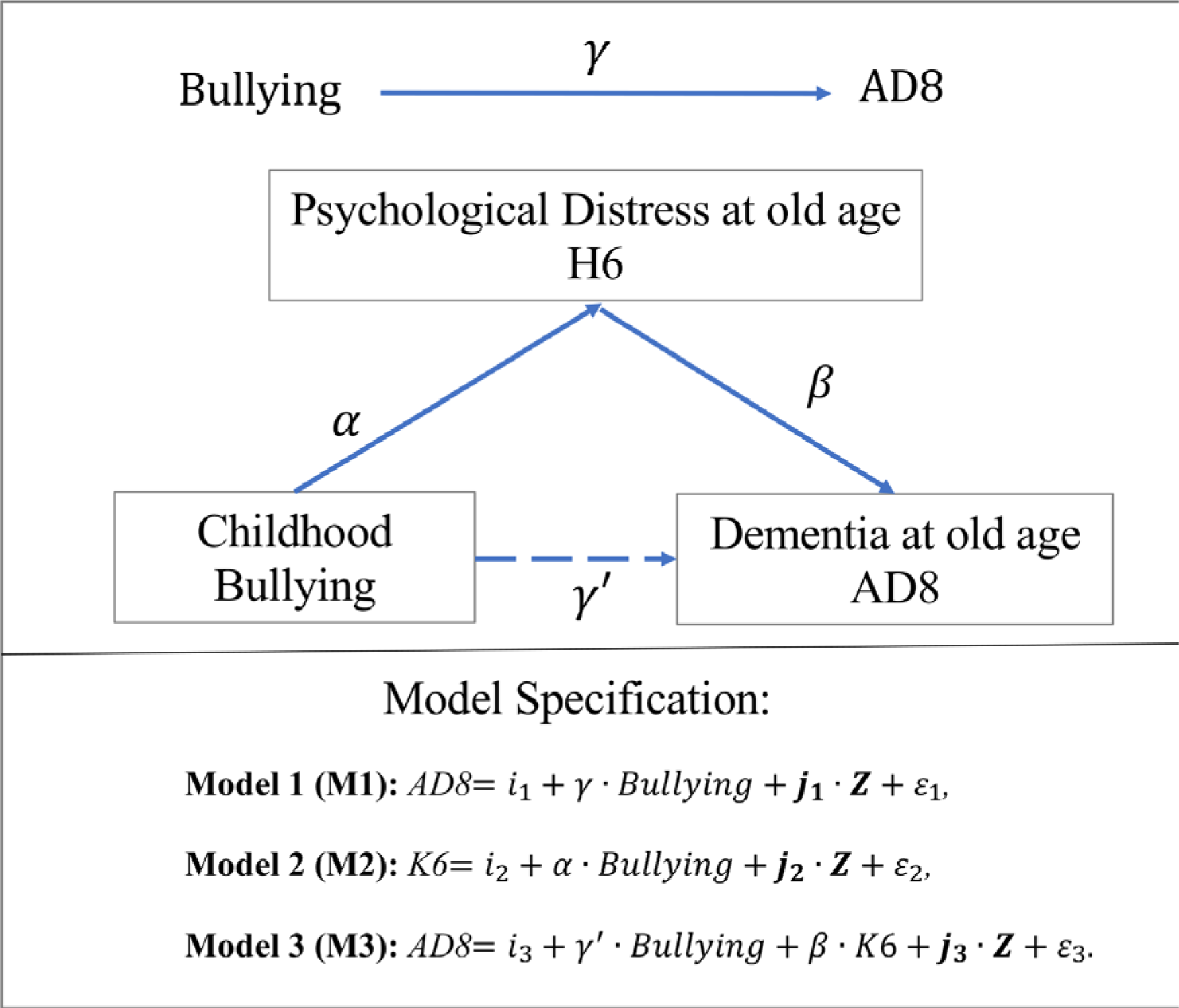
Schema of the mediation process for childhood bullying (6-16) and dementia risk later in life (65+). Models 1-3 represent the analytic approach behind this mediation model. In this model, **Z** is a vector of covariates that includes current age in 2017, sex, and the number of years of education. We assume the residual error terms (*ε*_1_, *ε*_2_, and *ε*_3_) are independent and normally distributed. We estimated parameters 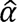, 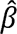, 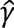, and 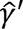 using Negative Binomial regression models.

For a better assessment of race differences and type of exposure, we conducted stratified models by race and type of bullying (target, bully, and bully-target). Sensitivity analysis additionally included models stratified by the age of exposure (ages 6-12 and 13-16). All models consider the complex survey design, and we report linearized standard errors for all estimates. We conducted all statistical analyses using Stata/SE© 16.1 (StataCorp LLC, College Station, TX).

## RESULTS

### Prevalence of childhood bullying experiences

**Figure 3** shows the distribution of Black and White respondents who experienced the three types of bullying experiences at ages 6 to 16 (bullies, targets, and those who were both a bully and a target). There is a higher percentage of White respondents who were targets of bullying compared to Black respondents (34.8% vs. 29.6%). There are also higher percentages of White respondents who were bullies (9.9% vs. 6.7%) and bully-targets (8.8% vs. 7.1%). When the data are disaggregated by age at exposure, we observe that a more significant percentage of White respondents (29.1%) were bullied at ages 6-12 compared to Black respondents (19.9%). In contrast, the percentages are very similar for White (21.1%) and Black respondents (20.6%) ages 13-16 (**Figure S1**).

**Figure 3.**
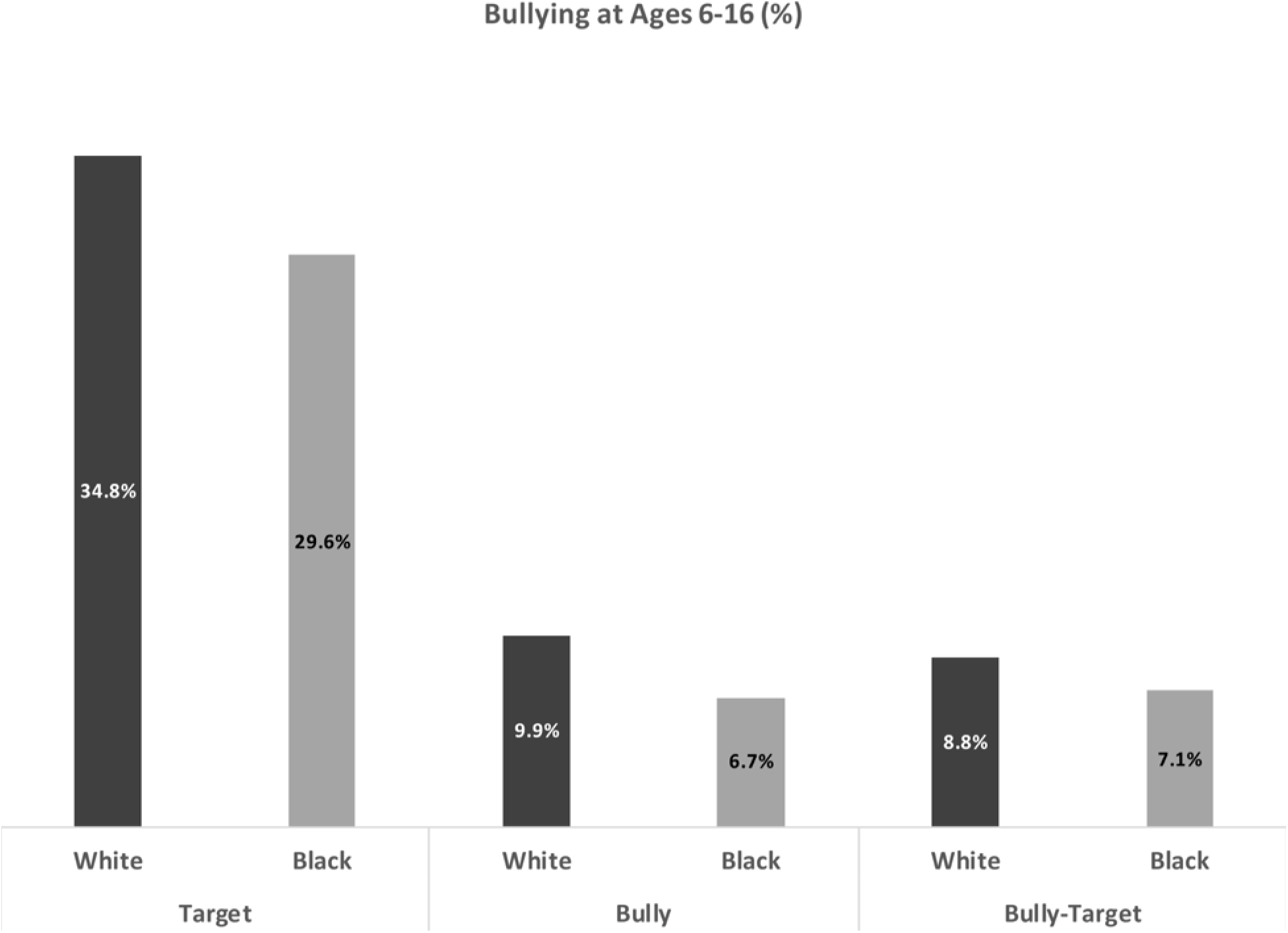
Percentage of individuals who were targets, bullies, or bully-targets during childhood by race. These estimates were determined for the final sample (523 respondents) described in Figure 2. In the case of Bully-Targets, we excluded those individuals who were only targets and only bullies.

### Model 1: Associations between childhood bullying experiences and late-life dementia risk

There is a significant association between childhood bullying experience and late-life dementia for Black respondents who were bullies during childhood/adolescence (Relative Risk [RR] = 3.03; 95% CI: 1.05, 8.69). However, we do not observe significant associations for Black respondents who were targets or bully-targets (**Table 2**). There are no significant associations between bullying (targets, bullies, and both) and late-life dementia for White respondents.

**TABLE 2.**
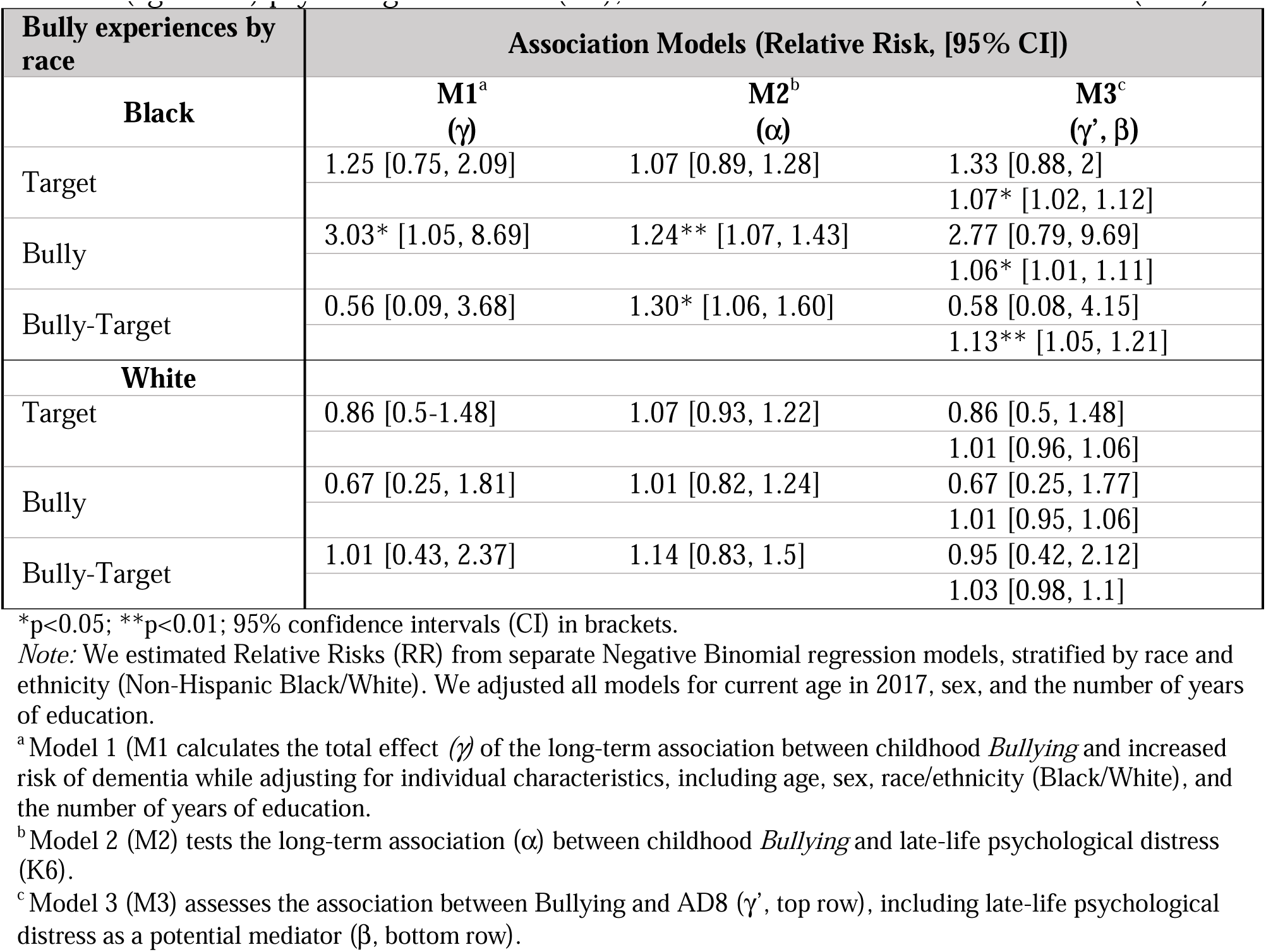
Long-term associations between childhood bullying (target, bully, and bully-target), late-life (aged 65+) psychological distress (H6), and late-life increased risk of dementia (AD8).

| Bully experiences by race | Association Models (Relative Risk, [95% CI]) |  |  |
| --- | --- | --- | --- |
| Black | M1<sup>a</sup><br>($\gamma$) | M2<sup>b</sup><br>($\alpha$) | M3<sup>c</sup><br>($\gamma'$, $\beta$) |
| Target | 1.25 [0.75, 2.09] | 1.07 [0.89, 1.28] | 1.33 [0.88, 2] |
|  |  |  | 1.07* [1.02, 1.12] |
| Bully | 3.03* [1.05, 8.69] | 1.24** [1.07, 1.43] | 2.77 [0.79, 9.69] |
|  |  |  | 1.06* [1.01, 1.11] |
| Bully-Target | 0.56 [0.09, 3.68] | 1.30* [1.06, 1.60] | 0.58 [0.08, 4.15] |
|  |  |  | 1.13** [1.05, 1.21] |
| White |  |  |  |
| Target | 0.86 [0.5-1.48] | 1.07 [0.93, 1.22] | 0.86 [0.5, 1.48] |
|  |  |  | 1.01 [0.96, 1.06] |
| Bully | 0.67 [0.25, 1.81] | 1.01 [0.82, 1.24] | 0.67 [0.25, 1.77] |
|  |  |  | 1.01 [0.95, 1.06] |
| Bully-Target | 1.01 [0.43, 2.37] | 1.14 [0.83, 1.5] | 0.95 [0.42, 2.12] |
|  |  |  | 1.03 [0.98, 1.1] |

\*p<0.05; \*\*p<0.01; 95% confidence intervals (CI) in brackets.
*Note:* We estimated Relative Risks (RR) from separate Negative Binomial regression models, stratified by race and ethnicity (Non-Hispanic Black/White). We adjusted all models for current age in 2017, sex, and the number of years of education.
<sup>a</sup> Model 1 (M1 calculates the total effect ( $\gamma$ ) of the long-term association between childhood *Bullying* and increased risk of dementia while adjusting for individual characteristics, including age, sex, race/ethnicity (Black/White), and the number of years of education.
<sup>b</sup> Model 2 (M2) tests the long-term association ( $\alpha$ ) between childhood *Bullying* and late-life psychological distress (K6).
<sup>c</sup> Model 3 (M3) assesses the association between Bullying and AD8 ( $\gamma'$ , top row), including late-life psychological distress as a potential mediator ( $\beta$ , bottom row).

### Model 2: Associations between childhood bullying experiences and late-life psychological distress

We note significant associations between bullying experience and late-life psychological distress among Black respondents who were bullies (RR = 1.24; 95% CI: 1.07, 1.43) or bully-targets (RR = 1.30; 95% CI: 1.06-1.60) (**Table 2**). There is not a significant association for Black respondents who were targets. We find no such associations among White respondents.

### Model 3: Associations between childhood bullying experiences and late-life dementia risk, mediated by late-life psychological distress

For White respondents, we observed that late-life psychological distress (i.e., answering in the affirmative for any of the K6 selections) strengthened the association between bullying experience and late-life dementia, but not enough to achieve significance (**Table 2**). In contrast, the associations for Black respondents do not show this same trend. Instead, all the M3 analyses of Black respondents show significant associations between late-life psychological distress and late-life dementia after including all bullying experiences.

### Comparison Across Models: Mediation Assessment

We next examined the impact of late-life psychological distress as a mediator of the association between bullying and dementia (i.e., M1 vs. M3). For Black respondents who were bullies, we observe that the association was attenuated from a RR of 3.03 in M1 to a non-significant RR of 2.77 (95% CI: 0.79, 9.69) in M3. The association of H6 with AD8 was positive and significant in M2 (RR = 1.24; 95% CI: 1.07, 1.43) and remained so in M3 after accounting for bullying experience (RR = 1.06; 95% CI: 1.01, 1.11) (**Table 2**). Given the non-significant association of bullying on AD8 in M3 (**Table 2**) and a significant indirect effect of H6 (i.e., psychological distress increases the association of bullying on dementia risk) (RR = 1.01; 95% CI: 1.00, 1.02) (**Table 3**) indicate evidence of full mediation (i.e., the association between childhood bullying and dementia risk is entirely explained by late-life psychological distress). In the case of bully-targets, while the association between psychological distress and the risk of dementia remained significant when considering the bullying experience (M2 vs. M3), we did not observe any indication of mediation effects (**Tables 2 and 3**).

**TABLE 3.** Direct and indirect effect associations derived from the full mediation model (M3) by type of bullying exposure (target, bully, and bully-target) and race (Black/White).

| Bully experiences by race | Direct ($\hat{\gamma}'$) and indirect ($\hat{\alpha}*\hat{\beta}$) estimates (Relative Risk, [95% CI]) | |
| --- | --- | --- |
|  | Black | White |
| Target: |  |  |
| Direct Effect | 1.32 [0.92-1.90] | 0.86 [0.51-1.43] |
| Indirect Effect | 1.00 [0.99-1.02] | 1.00 [0.99-1.00] |
| Bully: |  |  |
| Direct Effect | 2.77 [0.92-8.33] | 0.67 [0.26-1.68] |
| Indirect Effect | 1.01* [1.00-1.02] | 1.00 [0.999-1.001] |
| Bully-Target: |  |  |
| Direct Effect | 0.59 [0.10-3.28] | 0.95 [0.45-1.99] |
| Indirect Effect | 1.03* [1.00-1.06] | 1.00 [0.99-1.01] |

\*p<0.05; \*\*p<0.01.
*Note:* Relative Risks (RR) from direct and indirect associations from separate Negative Binomial regression models, stratified by race. We adjusted all models for current age in 2017, sex, and years of education. Direct associations correspond to the estimated parameter $\hat{\gamma}'$ in Model 3 (**Figure 3**). We evaluated indirect effects as the product of estimated parameters $\alpha$ from Model 2 and $\beta$ from Model 3 (**Figure 3**); that is: $\hat{\alpha}*\hat{\beta}$ .

### Sensitivity analysis

Our sensitivity analysis, which involved disaggregating models M1, M2, and M3 by the age of bullying exposure (childhood, ages 6-12, or adolescence, ages 13-16), revealed a significant influence of bullying during adolescence in the case of Black respondents who were bullies, with psychological distress partially explaining that relationship (**Table S1**). In the case of bully-targets, our findings revealed evidence of inconsistent mediation when the bullying experience occurred during adolescence. We observed significant associations for both direct and indirect effects; however, they were in oppositive directions. Several factors could contribute to this inconsistency, including the small sample size for this specific bullying experience, the potential influence of unaccounted confounding factors leading to bias or confounding effects, or the presence of suppression effects.^38^ Due to these limitations and uncertainties, our model cannot provide reliable evidence regarding the experience of Black respondents who were bully-targets. Finally, we did not observe significant associations for White respondents regardless of the age at which bullying occurred (**Table S1**).

## DISCUSSION

There is a significant gap in our understanding of how sociocultural and biobehavioral stresses experienced in early life contribute to risk of psychological distress and dementia in late life. The goal of this study was to test the hypothesis that exposure to childhood bullying increases the risk of dementia and late-life psychological distress for minority populations, specifically Black people, who may experience higher levels of early-life bullying and stress relative to Non-Hispanic White people.^20,39^ These effects may have long-lasting impacts over the life course. Using data from the Panel Survey for Income Dynamics (PSID), our results show that exposure to childhood bullying is associated with these deleterious long-term associations for Black respondents, but no such associations were observed for Non-Hispanic White respondents.

For Black respondents, those who were bullies had a higher risk of dementia after controlling for age, sex, and education. However, this association was completely explained by old-age psychological distress. In addition, late-life psychological distress was significantly associated with dementia risk in models that accounted for childhood bullying regardless of the type and age at exposure. In the case of bully-targets, we found no association between bullying and dementia, but psychological distress was associated with dementia regardless of the bullying experience. These results are consistent with previous literature indicating that late-life psychological distress, perceived stress, or depressive symptoms are associated with an increased risk of developing dementia.^40–43,22,25^

An interesting result is that these associations between bullying and dementia, mediated by late-life psychological distress, were only significant for Black respondents and not White respondents. Some studies provide a clue by reporting a higher or equivalent prevalence of depression among older Black residents compared to their White counterparts.^44^ Moreover, research reveals that Black adolescents who experience bullying are more prone to internalizing symptoms than perpetrators,^20^ and potentially carry considerable significance for developing late-life psychological distress.^45^ These associations were consistently significant when bullying occurred during adolescence (ages 13-16) rather than during childhood (ages 6-12).

Finally, it is worth emphasizing the significant association between being a bully with late-life psychological distress and the consistent association between late-life psychological distress and dementia risk among Black respondents. Previous studies have indicated that the long-term effects of childhood bullying on adulthood outcomes (ages 18-50) are contingent upon the specific type of bullying experienced. These studies have shown that bullied individuals experience worse outcomes, while those who solely acted as bullies exhibit few adverse effects. Bully targets, however, demonstrate the poorest outcomes regarding mental health and other adverse consequences.^46,47^ While this evidence aligns with our findings for White respondents who were bullies, it does not hold for Black respondents who were bullies. However, it is crucial to note that our results pertain to older adults aged 65 and above. These disparities in the long-term effects of bullying on adult health outcomes be attributed to how children and adolescents internalize or externalize problems based on their specific bullying experiences and across different racial and ethnic groups, as suggested by previous studies.^48,47^

Gaining a comprehensive understanding of the mediation effects of psychological distress across racial groups holds significant clinical importance. Such understanding enables physicians to potentially implement appropriate screening, preventive measures, and targeted interventions to mitigate the risk of dementia. Additionally, it emphasizes the need for culturally sensitive and tailored treatment approaches for individuals who have experienced childhood bullying. By recognizing the specific factors contributing to dementia development among different racial groups, healthcare practices, and policies can become more equitable. Furthermore, physicians can engage in meaningful discussions with patients about their experiences of childhood bullying, the potential long-term impact of psychological distress, and the association with dementia risk.

### Strengths and Limitations

This study presents a racially stratified analysis using early and late life stressors collected from the Panel Survey for Income Dynamics (PSID), which is the largest longitudinal panel study available in the world. This allowed us to test the separate and combined effects of bullying victimization and perpetration during childhood based on the age at which the exposure took place.

There are a few limitations to note. First, while the PSID is a comprehensive longitudinal panel dataset, the sample size for our analysis became relatively small when we restricted it to older adults and further disaggregated it by race. Second, this dataset lacks publicly available information about the participants’ other social stressors or any therapy they may have received during their lifetime, which may impact the perpetuation of stress later in adulthood. Third, the information about childhood bullying victimization is retrospective and subject to recall bias. Fourth, although K6 and AD8 are suitable proxy measures for psychological distress and dementia with good sensitivity and specificity, they are not clinical assessments. Fifth, the time interval between the measurement of H6 (2014) and AD8 (2017) falls within a 5-year interval (the standard set by other studies), but the exact onset of H6 or AD8 is unknown, then more research is needed to better understand the underlying mechanisms and the temporal relationship between psychological distress and dementia risk. Finally, this article does not comprehensively assess the mechanisms behind the indirect or direct effect associations found for Black and White respondents, but this may be the subject of future studies.

## CONCLUSION

This paper shows racial differences in the association between childhood bullying and late-life increased risk of dementia. The association was fully mediated by late-life psychological distress in the case of Black respondents who were bullies. This suggests that psychological distress could be a prodrome of dementia in this group. In the case of Non-Hispanic White respondents, however, we did not find any association or evidence of mediation. Further research is needed to examine the mechanisms behind the direct and indirect effect associations across racial/ethnic groups and the potential influence of other stressors during childhood. Gaining insights into the mediation effects of psychological distress among different racial groups is important in creating fair healthcare practices and policies. Such understanding not only facilitates the development of culturally sensitive and personalized treatment approaches but also addresses the needs of individuals who have endured childhood bullying.

## Data Availability

Scripts and datasets for this manuscript will be publicly available at Github (https://github.com/gauss75/BullyingDementiaPSID)

## ACKNOWLEDGMENTS

The collection of data used in this study was partly supported by the National Institutes of Health under grant number R01 HD069609 and R01 AG040213, and the National Science Foundation under award numbers SES 1157698 and 1623684.

## Author Contributions

IM-G conceived and designed the study. IM-G developed the analysis plan, conducted the quantitative analysis, and wrote the first draft of the manuscript. IM-G prepared the tables and figures. VSP contributed to writing the manuscript and provided critical insights. All authors contributed intellectually and gave final approval for publication.

## SUPPORTING INFORMATION

### Extended Data

**Figure S1.**
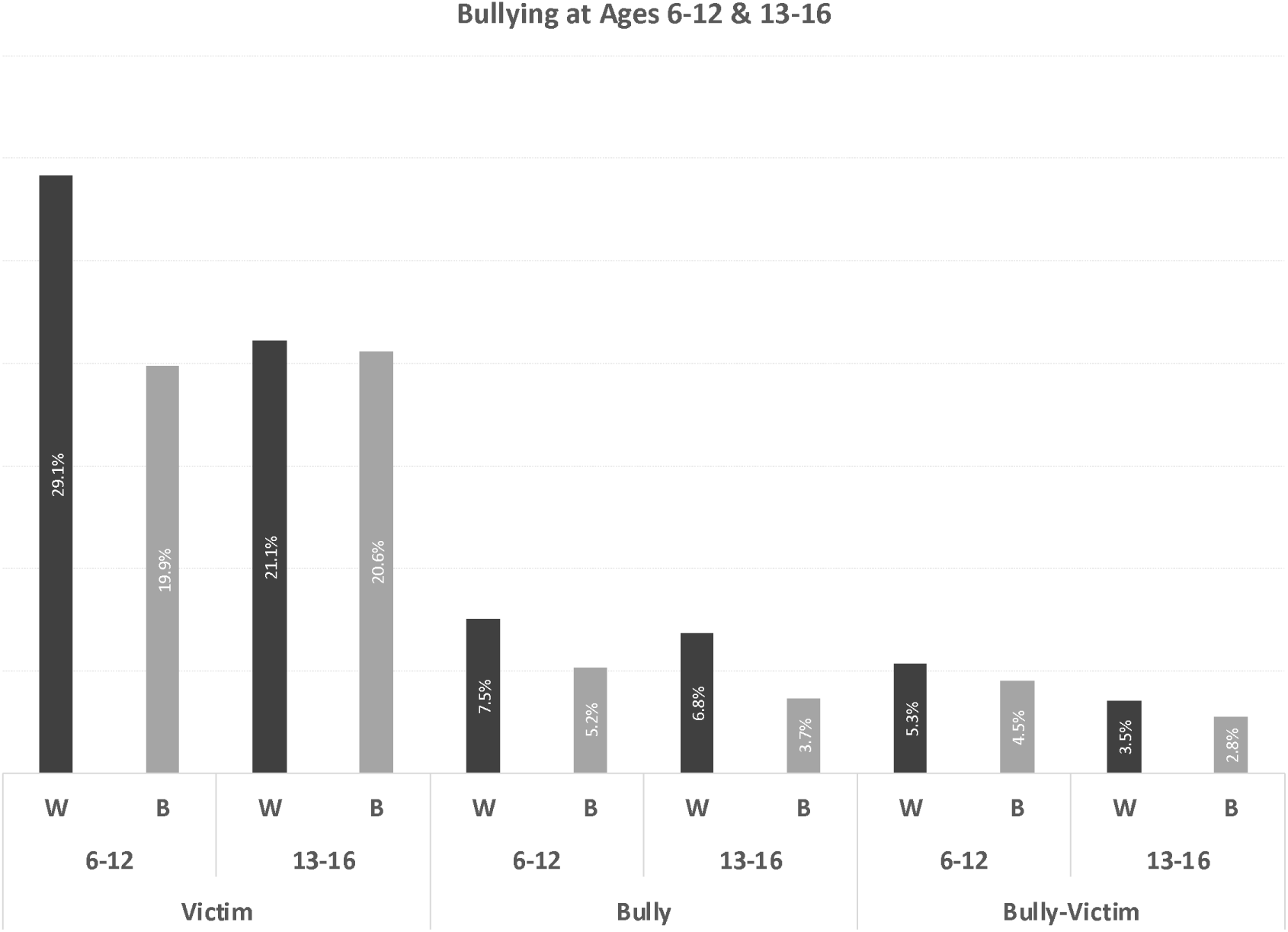
Percentage of targets, bullies, or bully-targets during childhood by race and age group. These estimates were determined for the final sample (523 respondents) described in Figure 2. In the case of Bully-Targets, we excluded those individuals who were only targets and only bullies from the reference group.

**Table S1.** Direct and indirect effect associations derived from the full mediation model (M3) by type of bullying exposure (target, bully, and bully-target), age group (6-12/13-16), and race (Black/White).

| Bully experiences by race | Direct ($\hat{\gamma}'$) and indirect ($\hat{\alpha}*\hat{\beta}$) estimates (Relative Risk, [95% CI]) | | | |
| --- | --- | --- | --- | --- |
|  | Black |  | White |  |
|  | Age 6-12 | Age 13-16 | Age 6-12 | Age 13-16 |
| Target: |  |  |  |  |
| Direct effect | 1.34 [0.97-1.83] | 0.68 [0.41-1.13] | 0.89 [0.47-1.7] | 0.86 [0.4-1.86] |
| Indirect Effect | 1 [0.99-1.01] | 1.01 [1-1.02] | 1 [1-1] | 1 [1-1] |
| Bully: |  |  |  |  |
| Direct effect | 0.63 [0.05-7.38] | 4.22** [2.24-7.97] | 0.53 [0.16-1.75] | 0.66 [0.19-2.3] |
| Indirect Effect | 1.01** [1-1.03] | 1.02** [1.01-1.04] | 1 [1-1] | 1 [1-1] |
| Bully-Target: |  |  |  |  |
| Direct effect | 0.81 [0.08-8.43] | 0.26** [0.15-0.47] | 1.07 [0.33-3.46] | 0.78 [0.24-2.47] |
| Indirect Effect | 1.02 [0.99-1.04] | 1.06** [1.01-1.1] | 1 [1-1.01] | 1 [0.99-1.01] |

